# Analytical sensibility and specificity of two RT-qPCR protocols for SARS-CoV-2 detection performed in an automated workflow

**DOI:** 10.1101/2020.03.07.20032326

**Authors:** Gustavo Barcelos Barra, Ticiane Henriques Santa Rita, Pedro Góes Mesquita, Rafael Henriques Jácomo, Lídia Freire Abdalla Nery

## Abstract

The World Health Organization declared that COVID-19 outbreak constituted a Public Health Emergency of International Concern and the development of reliable laboratory diagnosis of SARS-CoV-2 became mandatory to identify, isolate and provide optimized care for patients early. RT-qPCR testing of respiratory secretions is routinely used to detect causative viruses in acute respiratory infection. RT-qPCR in-house protocols to detect the SARS-CoV-2 have been described. Validations of these protocols are considered a key knowledge gap for COVID-19, especially if executed in a high throughput format. Here, we investigate the analytical sensitivity and specificity of two interim RT-qPCR protocols for the qualitative detection of SARS-CoV-2 executed in a fully automated platform. Under our conditions, the N1 and RdRP (modified) showed the highest analytical sensitivity for their RNA targets. E assay, in its original concentration, was considered a tertiary confirmatory assay. Taken together, N1, RdRP (optimized) and E presented appropriated analytical sensibility and specificity in our automated RT-qPCR workflow for COVID-19 virus, E being at least 4-fold less sensitive than the others. This study highlights the importance of local validation of in-house assays before its availability to the population. The use of the synthetic RT-qPCR target to investigate novel assays diagnostic parameters in automated workflows is a quick, simple effective way to be prepared for upcoming threats. The proposed assay detected the fisrt SARS-CoV-2 infection in Brazilian Central-West.

## Introduction

On 30 January 2020, the World Health Organization declared that the COVID-19 outbreak constituted a Public Health Emergency of International Concern and the development of reliable laboratory diagnosis of SARS-CoV-2 became mandatory to identify, isolate and provide optimized care for patients early^1^. RT-qPCR testing of respiratory secretions is routinely used to detect causative viruses in acute respiratory infection and, during a Public Health Emergency of International Concern, the establishment of standardized processes and protocols, as well as sharing of specimens, data, and information is critical. RT-qPCR in-house protocols to detect the SARS-CoV-2 have been described^2^. Validations of these protocols are considered a key knowledge gap for COVID-19, especially if executed in a high throughput format. Here, we investigate the analytical sensitivity and specificity of two interim RT-qPCR protocols^3,4^ for the qualitative detection of SARS-CoV-2 executed in a fully automated platform.

## Methods

### Samples and collection tubes

Sixty nasopharyngeal swabs samples were collected from healthy volunteers using Rayon swab and placed into tubes containing 4.3 mL of guanidine hydrochloride (40%), which virtually inactivates the virus and preserves all RNA in the specimen.

### Ethical considerations

All volunteers agreed to participate, signed informed consent and the internal use of these samples for diagnostic workflow optimization was according to the medical ethical rules of our institution.

### SARS-CoV-2 RT-qPCR protocols

CDC and Charité described the interim SARS-CoV-2 RT-qPCR protocols evaluated in this study. Assays names described by each protocol were maintained to avoid mistakes: N1, N2, N3, N, E, RdRP. Primers and probes for SARS-CoV-2 can be found in the supplemental file.

### Collection and process RT-qPCR quality control

Each viral primer/probe set was multiplexed with primer/probe set for human RPP30 gene to control the sample collection, as the amount of biological material varies from sample to sample, and with a primer/probe set for an artificial RNA sequence (artificial external control - AEC), which is introduced during extraction, to control the RT-qPCR steps and its inhibition. Primers and probes for RPP30 and AEC can be found in the supplemental file.

### RT-qPCR workflow

The RT-qPCR workflow was executed on Flow Flex Solution, which is composed by a liquid handlers and a nucleic acid extractor. The liquid handler pipettes the primary samples and distribute the PCR reaction master mix. Nucleic acids were extracted from 200 ul of the primary sample using MagNA pure 96 DNA and Viral NA Small Volume Kit on Magna pure 96 instrument. One-step RT-qPCR reaction was prepared with LightCycler® Multiplex RNA Virus Master (all from Roche Diagnostics, Mannheim, Germany). Primers/probe concentrations suggested by the interim RT-qPCR protocols providers were maintained and other concentrations were also tested (table 1). The final reaction volume was 10 ul and the automated workflow was able to perform 4 viral assays in 96 samples in 4-5 hours.

**Table 1.** Primer and probes concentration used in this study

| **Primer** | **N1** | **N2** | **N3** | **RpDP** | **RpDP Modified** | **E** | **E Modified** | **N** |
| --- | --- | --- | --- | --- | --- | --- | --- | --- |
| Primer F | 1500 nM | 1500 nM | 1500 nM | 600 nM | 1500 nM | 400 nM | 1500 nM | 600 nM |
| Primer R | 1500 nM | 1500 nM | 1500 nM | 800 nM | 1500 nM | 400 nM | 1500 nM | 800 nM |
| Probe 1 | 375 nM | 375 nM | 375 nM | 100 nM | 375 nM | 200nM | 375 nM | 200 nM |
| Probe 2 | - | - | - | 100 nM | - | - | - | - |

## Results

All nasopharyngeal swabs samples from healthy volunteers were submitted to the six considered SARS-CoV-2 assays to check the generation of unspecific response. We observed consistent false-positive results for N and N2 (60 out of 60 samples for both). N3 assay generated false positive signal or inconclusive results in 13 out of 60 tested samples. N2, N3, and N assays were not considered for subsequent experiments due to the lack of analytical specificity under our conditions (figure 1).

**Figure 1.**
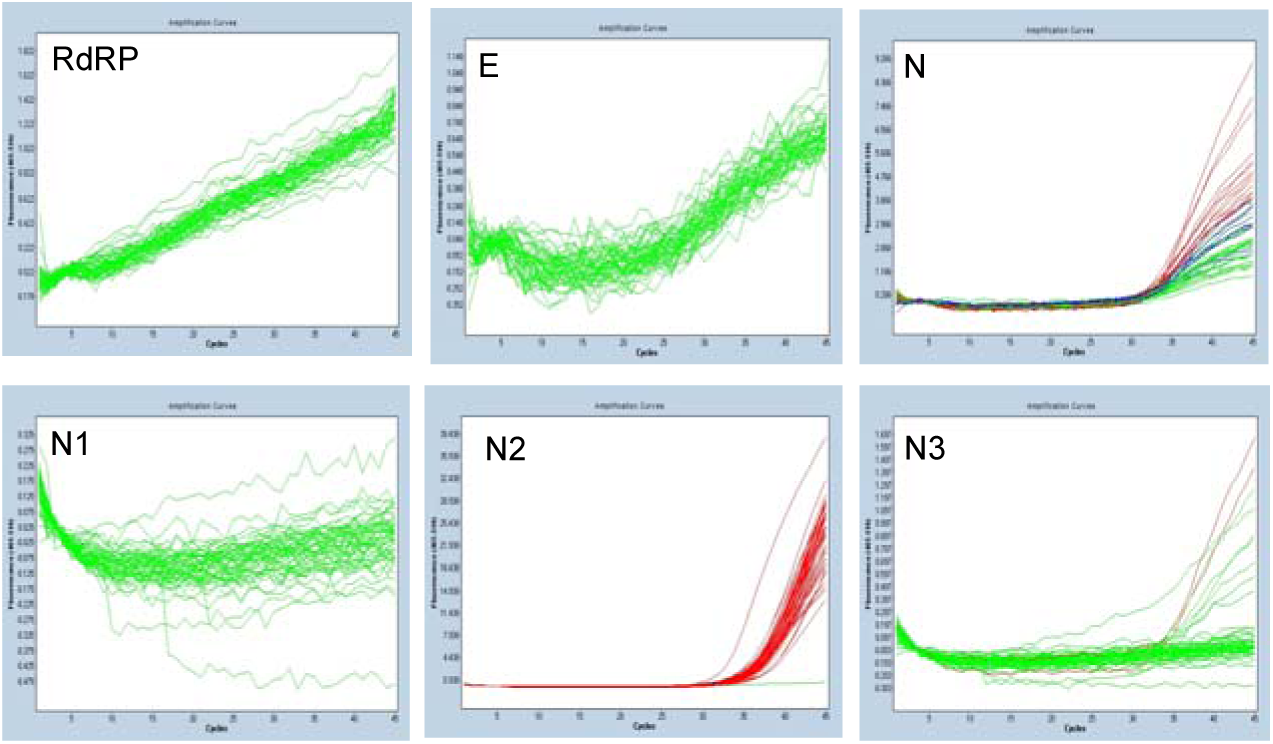
Assays analytical specificity analysis. N, N2, and N3 were excluded from the validation due to lack of analytical specificity when applied to healthy volunteer samples.

A synthetic dsDNA molecule (Integrated DNA Technologies, Coralville, USA) comprising of the concatenation of all six viral assays target sequences in the SARS-CoV-2 genome was *in vitro* transcribed into its RNA form. This SARS-CoV-2 diagnostic synthetic RNA was quantified and a previously known copy number solution was spiked into negative swabs to construct positive samples as similar as possible from real clinical samples. These constructed samples were submitted to the automated workflow for assays that passed in the analytical specificity evaluation (N1, E, and RdRP).

Limiting dilution of 1:2 (from 2.64×10^4^ to 4.04×10^−1^ copies/reaction) of the synthetic SARS-CoV-2 diagnostic RNA was tested in order to evaluate the assays’ limit of detection. Probit regression analysis returned the limit of detection of 21 (95% CI 16.5 - 31.1) copies/reaction for N1, 141 (95% CI 109 - 207) copies/reaction for E and 350 (95% CI 281 - 508) copies/reaction for RdRP (Figure 1, B). E and RdRP assay primer/probe concentrations were modified and limits of detection changed to 457 (95%CI 382 - 598) and 33.7 (95% CI 27.6 – 46.8) copies/reaction, respectively. Primers/probe concentration optimization did not interfere with specificity of the assays (20 healthy volunteers samples tested).

Amplifications efficiencies of N1, RdRP, RdRP (modified), E, and E (modified) assays were 93.4%, 116.5%, 110%, 86% and 119.6%, respectively, when applied to serial dilution of 1:10 (1.48×10^8^ to 1.48×10^2^ copies/PCR) of the synthetic SARS-CoV-2 diagnostic RNA. N1 and RdRP (modified) showed better amplification efficiency and this observation corroborates with the results of the limiting dilution experiment because assays with amplification efficiencies closed to 100% have better diagnostic performance (figure 3).

**Figure 2.**
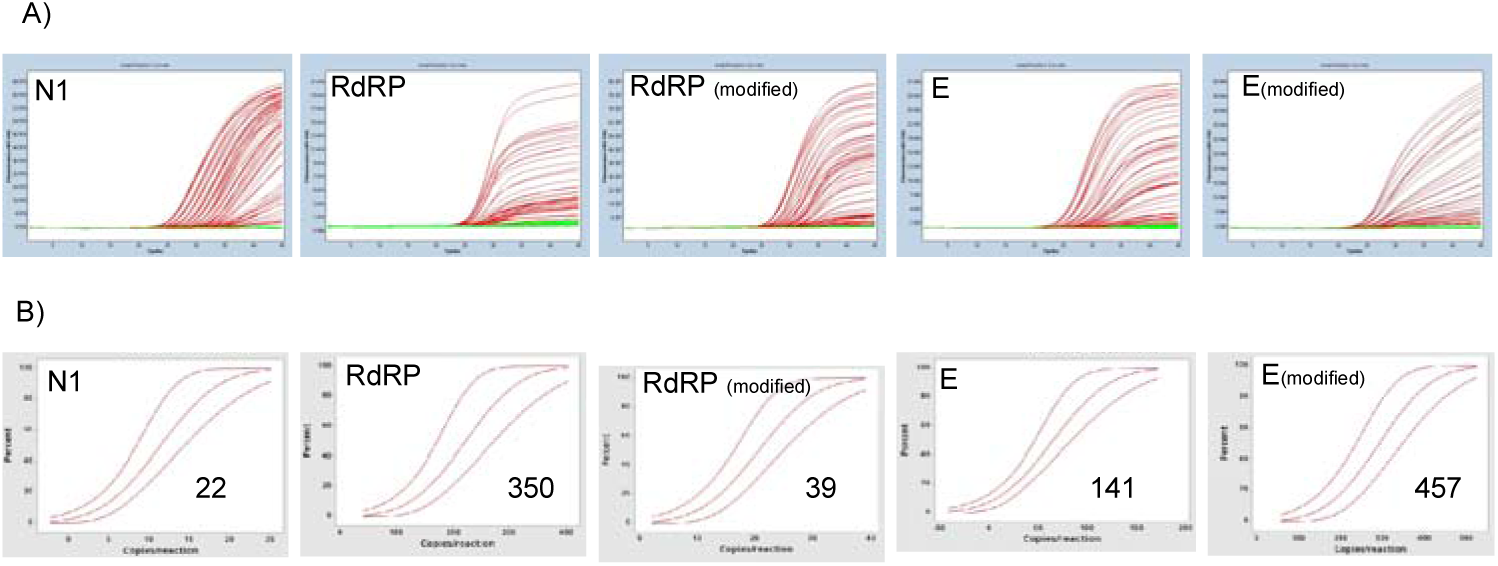
Assays limit of detection determination. N1 and RdRP (modified) showed better LOD. A) Raw data and B) Probit regression analysis (inserted unit values are copies/reaction).

**Figure 3.**
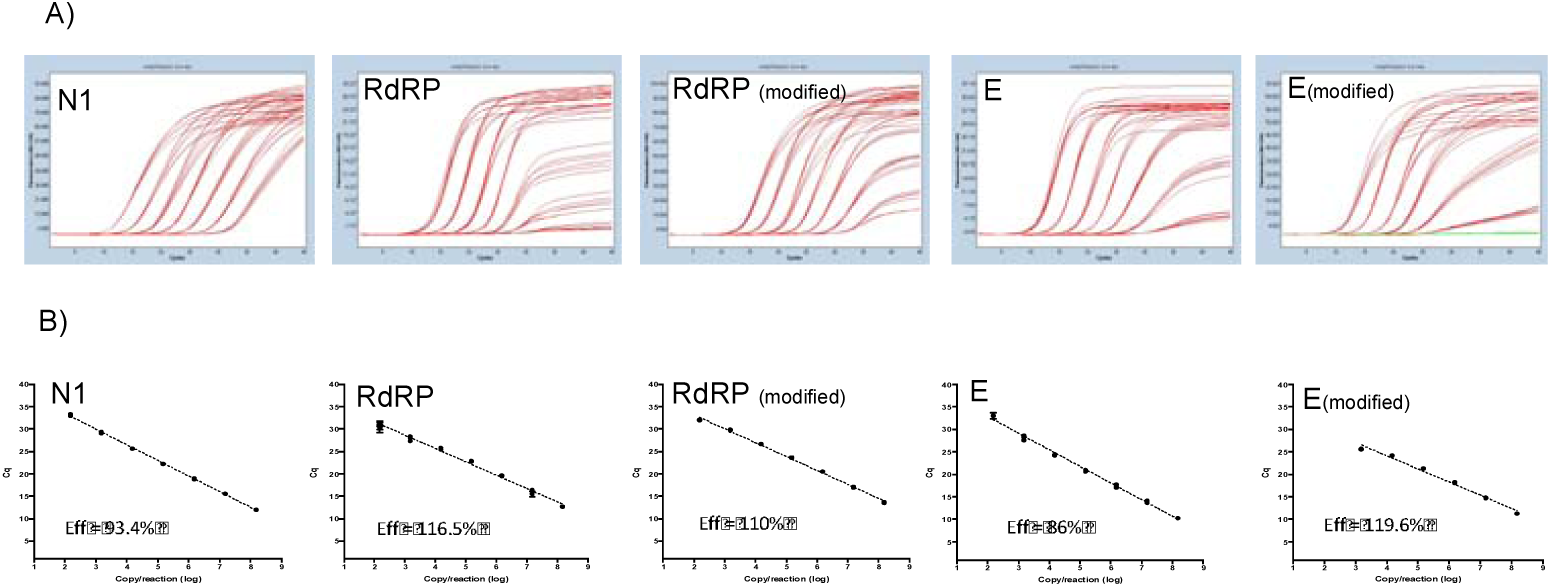
Assays amplification efficiency analysis. N1 and RdRP (modified) showed better amplification efficiencies. A) Raw data and B) Regression analysis.

When applied to pools of positive samples used in our laboratory as internal quality controls for RT-qPCR, assays for other respiratory viruses were tested (Influenza A H1N1 and H3N2, parainfluenza virus 3 and 4, rhinovirus, coronavirus 229, coronavirus HKU) N1, RdRP, RdRP (modified), E, and E (modified) assays did not returned false-positive results.

## Discussion

So, under our conditions, the N1 and RdRP (modified) showed the highest analytical sensitivity for their RNA targets. E assay, in its original concentration, was considered a tertiary confirmatory assay. Taken together, N1, RdRP (optimized) and E presented appropriated analytical sensibility and specificity in our automated RT-qPCR workflow for COVID-19 virus, E being at least 4-fold less sensitive than the others.

The sensitivities observed in this study were slightly different than the described for RdRP (3.6 copies per reaction) and E (3.9 copies per reaction) original description, where the authors used the *in vitro* transcribed SARS-CoV-2 RNA directly in the reaction^3^. Our results can be secondary to the fact that we spiked the synthetic RNA in the nasopharyngeal samples, resembling a real clinical sample and in the limit of detection calculation, we assumed that the nucleic acid purification recovered 100% of the spiked RNA sequences of the 200 ul aliquot used in the extraction.

This study highlights the importance of local validation of in-house assays before its availability to the population. Experiments to establish the assay analytical specificity can be easily implemented. At this moment, the proposed method [N1 (primary) and RPDP (modified) (confirmatory)] detected the first case of SARS-CoV-2 Brazilian central-west region, demonstrating that the use of the synthetic RT-qPCR target to investigate novel assays diagnostic parameters in automated workflows is a quick, simple, effective way to be prepared for upcoming threats. The use of spiked samples resembling a real clinical specimen exposes the artificial SARS-CoV-2 sequences to the same background of nucleic acids yields that can be found in the routine and similar amplification behavior of a real SARS-CoV-2 is expected.

## Data Availability

Not applied

